# Pace of Aging in older adults matters for healthspan and lifespan

**DOI:** 10.1101/2024.04.25.24306359

**Authors:** A Balachandran, H Pei, Y Shi, J Beard, A Caspi, A Cohen, BW Domingue, Indik C Eckstein, L Ferrucci, A Furuya, M Kothari, TE Moffitt, C Ryan, V Skirbekk, Y Zhang, DW Belsky

**Affiliations:** Robert N Butler Columbia Aging Center, Columbia University Mailman School of Public Health, New York, NY, USA; Department of Epidemiology, Columbia University Mailman School of Public Health, New York, NY, USA; Department of Health Policy & Management, Columbia University Mailman School of Public Health, New York, NY, USA; Department of Psychology & Neuroscience, Duke University, Durham, NC, USA; Social, Genetic, and Developmental Psychiatry Unit, Institute of Psychiatry, Kings College, University of London, London, UK; Department of Environmental Health Sciences, Columbia University Mailman School of Public Health, New York, NY, USA; Graduate School of Education, Stanford University, Palo Alto, CA, USA; National Institute on Aging, Bethesda, MD, USA; ICAP, Columbia University Mailman School of Public Health, New York, NY, USA; Norwegian Institute for Public Health, Oslo, Norway; Department of Sociomedical Sciences, Columbia University Mailman School of Public Health, New York, NY, USA

## Abstract

As societies age, policy makers need tools to understand how demographic aging will affect population health and to develop programs to increase healthspan. The current metrics used for policy analysis do not distinguish differences caused by early-life factors, such as prenatal care and nutrition, from those caused by ongoing changes in people’s bodies due to aging. Here we introduce an adapted Pace of Aging method designed to quantify differences between individuals and populations in the speed of aging-related health declines. The adapted Pace of Aging method, implemented in data from the US Health and Retirement Study and English Longitudinal Study of Aging (N=21,463), integrates longitudinal data on blood biomarkers, physical measurements, and functional tests. It reveals stark differences in rates of aging between population subgroups and demonstrates strong and consistent prospective associations with incident morbidity, disability, and mortality. Pace of Aging can advance the population science of healthy longevity.

## INTRODUCTION

Population aging is the central demographic phenomenon of the century and is unprecedented in human history^1,2^. Current trends are driven primarily by gains in life expectancy for older adults^3–5^. In some places and population subgroups, these gains in life expectancy have been matched by gains in healthy years of life (“healthspan”)^6,7^. However, in other cases, healthspan and lifespan are diverging, with potential to disrupt healthcare systems and economies^3,8,9^. The current toolkit available to demographers and public health planners allows differentiation of populations with more and less successful aging only in terms of completed lifespans or healthspans ^10–12^. These metrics are effective in summarizing differences between populations accumulated across the full life course. However, they do not distinguish deficits in health established early in life from the ongoing changes in people’s bodies that are the essence of aging^13^. Both are important. However, only the latter are expected to respond to healthy-longevity interventions with midlife and older adults^14^. As a consequence, the metrics in our existing toolkit may not be adequately sensitive to the effects of such interventions.

Measurements are needed that can distinguish ongoing aging-related changes in organs, tissues, and capacities from differences in health that are legacies of early life in order to provide optimally-sensitive metrics for population surveillance and intervention evaluation. We previously developed the Pace of Aging method to quantify individual differences in aging trajectories with the goal of informing design and evaluation of clinical interventions ^15,16^. Here we adapt this methodology with the goal of informing evaluations programs and policies for aging societies.

Our original Pace of Aging method was developed from analysis of health changes from young adulthood to midlife in the Dunedin Study 1972-73 birth cohort^15^. To be most useful for comparative biodemographic analysis used by planners to evaluate efforts to promote healthy longevity, the Pace of Aging method needs to be adapted to a different context: samples of individuals representing a wide range of birth cohorts for whom follow-up begins later in the life course. In addition, whereas the Dunedin Study collected extensive biochemical and physical examination data from participants, the studies used by planners typically have access to much sparser measurement panels.

Here, we introduce an adapted method for calculation of Pace of Aging in a sample composed of a wide range of birth cohorts with follow-up in midlife and older age and a sparse panel of biomarkers. Our adapted method is designed to generalize across the Gateway to Global Aging family of harmonized cohort studies^17^, which comprises cohorts across Europe, Asia, and Central and South America that are routinely cited in planning and policy analysis. We implement the method and test proof of concept using data from the US Health and Retirement Study (HRS). As the other cohorts extend their biomarker follow-up, the method introduced here can be applied to conduct cross-national comparative analysis.

We compiled data from dried-blood spot, physical exam, and functional test protocols conducted by the HRS during 2006-2016 (six assessment waves). We identified nine parameters measured at all six waves that met criteria for inclusion in the Pace of Aging analysis: C-reactive protein (CRP), Cystatin-C, glycated hemoglobin (HbA1C), diastolic blood pressure, waist circumference, lung capacity (peak flow), tandem balance, grip strength, and gait speed. A total of 13,573 individuals provided data on at least six of these nine biomarkers across at least two of the follow-up assessments. We modeled longitudinal change in these biomarkers to estimate person-specific slopes for each of them. Then we combined slope information across biomarkers to compute each participants’ Pace of Aging. Our validation analyses tested associations of Pace of Aging with measures of morbidity, disability, and survival through 2021. Finally, we evaluated socioeconomic and demographic disparities in the Pace of Aging among US older adults. We conducted parallel analysis to develop and validate a Pace of Aging phenotype in the English Longitudinal Study of Aging (ELSA).

## METHODS

### Sample

The Health and Retirement Study (HRS) is a nationally representative longitudinal survey of US residents ≥50 years of age and their spouses. The HRS has been fielded every two years since 1992. Participants are asked about four broad areas: income and wealth; health, cognition, and use of healthcare services; work and retirement; and family connections. A new cohort of 51– 56-year-olds and their spouses is enrolled every six years to maintain representativeness of the U.S. population over 50 years of age. Response rates over all waves of the HRS range from 81-91%. As of the most recent data release, HRS included data collected from 42,515 individuals in 26,600 households. We linked HRS data curated by RAND Corporation^18^ with dried-blood-spot biomarker data collected during 2006-2016^19^.

English Longitudinal Study of Aging (ELSA) is a nationally representative longitudinal survey of residents ≥50 years of age and their cohabitating spouses in private households of England. ELSA has been fielded every two years since 2002-2003. ELSA was modelled after the HRS in the US, and asked participants about their health, economic position, and quality of life. New cohorts of age >50 was added from Wave 3 (2006-07) to maintain representativeness of the English population over 50 years of age. Response rates of the ELSA ranges 55-82%. with the original sample consisting of 11,391 individuals. We linked ELSA data curated by USC Gateway to Global Aging (Reference added in Zotero) with blood -biomarker data collected during nurse home visits to participants over the period 2004-2012 (Reference).

### Measures

#### Pace of Aging

We measured Pace of Aging from blood biomarker, physical assessment, and functional test performance data collected during home visits to participants during 2006-2016. Biomarkers were included in Pace of Aging analysis based on known connections with processes of aging and expectation of monotonic change across the age range included in the HRS. Three blood biomarkers (HbA1c, C-reactive protein, cystatin-C), three physical assessments (diastolic blood pressure, peak-flow lung-function testing, waist circumference), and three functional tests (gait speed, balance, grip strength) were included in analysis. (We selected diastolic blood pressure as the blood pressure parameter for Pace of Aging analysis because it is expected to decline with aging and therefore its expected trajectory would not be reversed by antihypertensive therapy. We excluded three lipid measures, total cholesterol, high-density lipoprotein cholesterol, and low-density lipoprotein cholesterol, because these biomarkers are under routine medical management in older people and are known to exhibit nonlinear changes with aging across the age-range of our cohorts.) Participants received home visits at 4-year intervals (2006, 2010, and 2014 or 2008, 2012 and 2016). We measured Pace of Aging using data from participants with at least 2 repeated measures of at least six of the nine biomarkers representing all three types of data and who were younger than age 90 at the time of their first biomarker measurement (N=13,573). Baseline observations were recorded in 2006 for 39% of the sample, in 2008 for 34%, in 2010 for 14% and in 2012 for 13%. Three repeated measures were available for 55% of the sample. Characteristics of each of these groups of participants are reported in **Supplemental Table 1**. Biomarker measurements are described in detail in **Supplemental Table 2** and **Supplemental Figure 1**. Biomarker summary statistics are reported in **Supplemental Table 3**.

We modeled Pace of Aging in 4 steps. First, we standardized biomarkers to a common distribution by centering values on the sex-specific mean for participants aged <65 years of age (the mean age of participants in our sample) and dividing the centered values by the sex-specific standard deviation of that group (Gait speed was measured in participants aged 65 and older; we standardized values based on distributions for participants aged 65-75). Age-group specific means and standard deviations for men and women included in the analysis are reporting in **Supplemental Table 3**. For biomarkers that decline with aging (diastolic blood pressure, peak flow, balance, and grip strength), standardized values were reversed so that higher values corresponded to poorer organ-system function. Gait speed was measured as the natural log of time to complete the walk test and so did not need to be reversed.

Second, we modeled change over time in each biomarker using linear mixed-effects regression models with participant-specific random intercepts and slopes. Models included baseline year of biomarker measurement, follow-up time, chronological age at baseline (modeled as a 3^rd^ degree B-spline) and an interaction between age at baseline and follow-up time as covariates. Interactions between baseline-age terms and time were included to allow for variation in slopes of change depending on participants’ ages at baseline. Models were fitted separately for men and women.

Third, for each biomarker, we measured pace of change for each participant by combining fixed and random slope components.

Finally, we computed each participant’s Pace of Aging as the average pace of change across the nine biomarkers. We scaled Pace of Aging based on the sex-specific average of participants under age 65. For the resulting measure, a value of 1 represents average biomarker change per chronological year in HRS participants aged <65. A value of 1.5 would indicate a Pace of Aging 50% faster relative to this average. A value of 0.5 would indicate a Pace of Aging 50% slower relative to the average.

We followed a parallel analysis plan to develop a Pace of Aging measurement in ELSA. Details are reported in **Figure 1** and **Supplemental Tables 2 and 4**, and **Supplemental Figure 1**.

**Figure 1.**
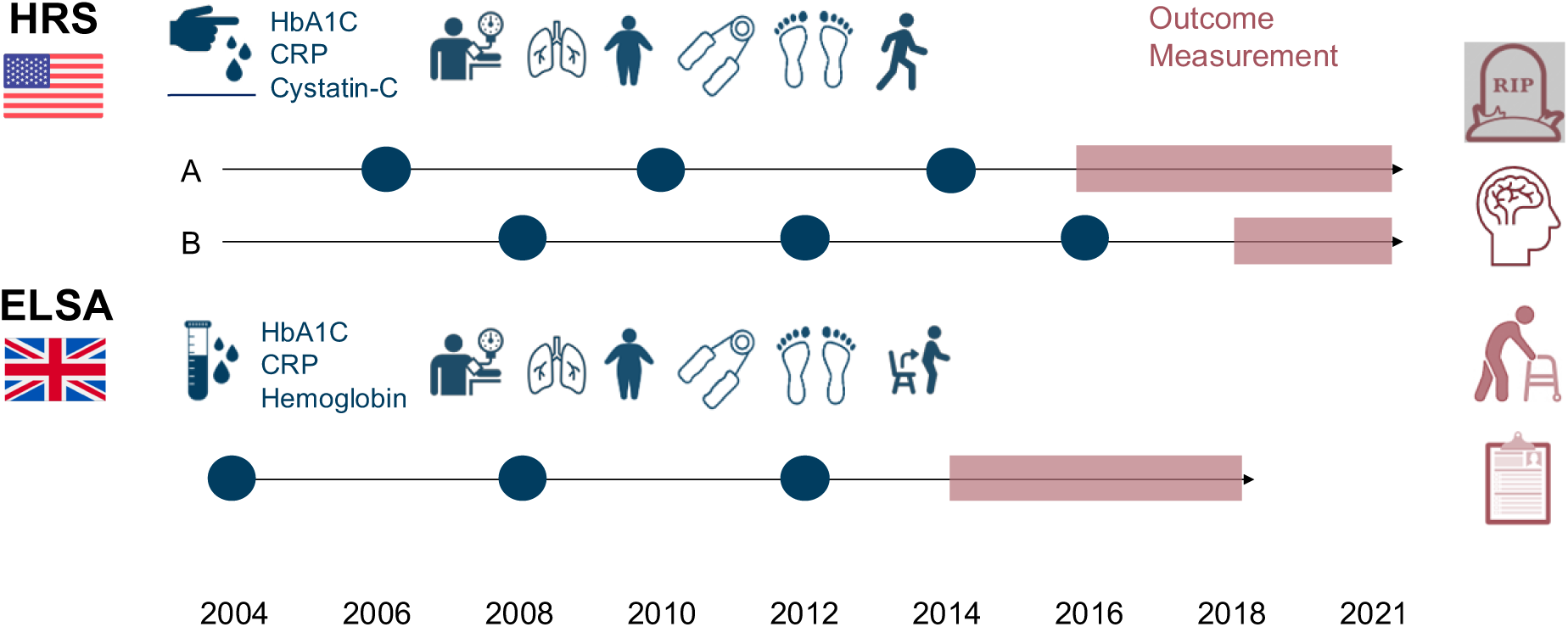
Study Design for Pace of Aging analysis in the US Health and Retirement Study and the English Longitudinal Study of Aging. Figure shows design of data collection in the US Health and Retirement Study (HRS, N=13,573) and the English Longitudinal Study of Aging (ELSA, N=7,890) for Pace of Aging analysis. Measurements included in Pace of Aging are illustrated above study timelines for HRS (top) and ELSA (bottom). Measurements of healthspan outcomes are illustrated on the right side of the figure. Timing of collection of Pace of Aging measurements is shown in blue circles. For HRS, measurements were collected on two different schedules, each including roughly half of the cohort. For ELSA, all measurements were taken on the same schedule. Timing of collection of healthspan outcomes is shown in shaded red bars (2016-2021 and 2018-2021 for HRS; 2014-2018 for ELSA).

#### Mortality, morbidity, disability and cognitive impairment

We evaluated criterion validity of the Pace of Aging by testing associations with four components of healthy lifespan: survival, incidence of chronic disease, incidence of disability and decline in cognition. Survival was measured by HRS through the 2020 follow-up assessment. We analyzed survival from the baseline biomarker measurement. Chronic disease incidence was measured from participant reports of physician diagnosed chronic disease. Disability incidence was measured from participant reports about Activities of Daily Living (ADL) limitations and Instrumental Activity of Daily Living (IADL) limitations. Cognitive decline and incident cognitive impairment were measured according to the Langa-Weir approach ^21^ from cognitive-test and interview data.

Survival, chronic disease, disability, and cognition measures are described in detail in **Supplemental Table 4**. All outcomes other than mortality were measured at the 2020 measurement wave or the last wave a participant contributed data subsequent to the end of follow-up for pace of aging measurement.

The ELSA study has so far released data only through 2018 and not yet released data files with measurements of survival and dementia classification. Therefore, the parallel measurement battery in ELSA included only chronic disease incidence, ADLs, IADLs, and cognitive test performance through 2018. Details are reported in **Supplemental Table 4.**

### Analysis

We used regression analysis to test Pace of Aging associations with healthspan phenotypes. We fit Cox proportional hazard models to estimate hazard ratios (HRs) and 95% CIs for mortality. We fit Poisson regression models to estimate incidence rate ratios (IRRs) and 95% confidence intervals (CIs) for incident chronic disease and limitations to activities of daily living (ADLs) and instrumental activities of daily living (IADLs). We use multinomial logit models to estimate odds-ratios (OR) for incident cognitive impairment and dementia. We use linear regression models to estimate change in cognitive-function scores between baseline and follow-up and to test differences in Pace of Aging between population sub-groups. Models included participants aged 50-90 years with measured Pace of Aging and the outcome or exposure of interest (maximum HRS N=13,317; maximum ELSA N=7,890). All models were adjusted for sex, race/ethnicity, age, age-squared, and a set of terms encoding baseline measurement year and whether the participant contributed two or three repeated measures of biomarker data. Models of healthspan outcomes other than mortality included an offset variable for follow-up time.

## RESULTS

We analyzed data from US Health and Retirement Study (HRS) participants aged 40 or older at the time of their first biomarker measurement who contributed at least two repeated measures of six or more biomarkers over 2006-2016 (N=13,573 41% male, mean age at baseline=65, SD=10). This analysis sample was slightly younger and better educated in comparison to the overall HRS sample (**Supplemental Table 1**). Baseline observations were recorded in 2006 for 39% of the sample, in 2008 for 34%, in 2010 for 14% and in 2012 for 13%. Three repeated measures were available for 55% of the sample. Summary statistics for biomarker measurements at baseline are reported in **Supplemental Table 3**.

### Older adults showed signs of correlated decline in multiple indicators of system integrity over 4-8 years of follow-up

Of the nine biomarkers included in HRS analysis, eight showed the expected pattern of change: Gait speed, grip strength, balance, diastolic blood pressure, and peak-flow declined; cystatin-C, HbA1c, and waist circumference increased. For CRP, change was in the expected positive direction for men, but declined slightly for women. Slopes of aging-related decline were steepest for the functional test biomarkers. Results were similar in ELSA. Biomarker slopes of change are reported in **Supplemental Table 5** and are plotted in **Supplemental Figure 2.**

Participants who were older at baseline showed steeper slopes of change in most biomarkers. Exceptions were HbA1c and waist circumference, possibly reflecting declining weight gain in later life. Slopes of change were positively correlated across biomarkers, again with the exceptions of HbA1c and waist circumference. Correlations among slopes of change are reported in **Supplemental Figure 3**. In the next step, we composited slopes of change across biomarkers to quantify the overall rate of decline in system integrity.

### Pace of Aging

We computed each participant’s Pace of Aging as the average pace of change across the nine biomarkers. We scaled Pace of Aging based on the sex-specific average value for participants under age 65. Resulting values can be interpreted as years of biological change per calendar year relative to the reference group. HRS Pace of Aging values were approximately normally distributed and indicated faster aging in men as compared to women and older as compared to younger participants (Pace of Aging mean=1.33 (SD=0.68); correlation with chronological age at baseline r=0.61; male-female difference Cohen’s d=0.21, 95% CI [0.19-0.24]). We followed the same procedure in ELSA. Results were similar, although correlation with chronological age was somewhat stronger and sex differences were smaller. Age and sex differences in Pace of Aging values are illustrated in **Supplemental Figure 4.** Contributions of slopes within each of the three biomarker categories to Pace of Aging and intercorrelations of slopes across biomarker categories are shown in **Supplemental Figure 5**. Individual participant Pace of Aging slopes are plotted in **Figure 2**.

**Figure 2.**
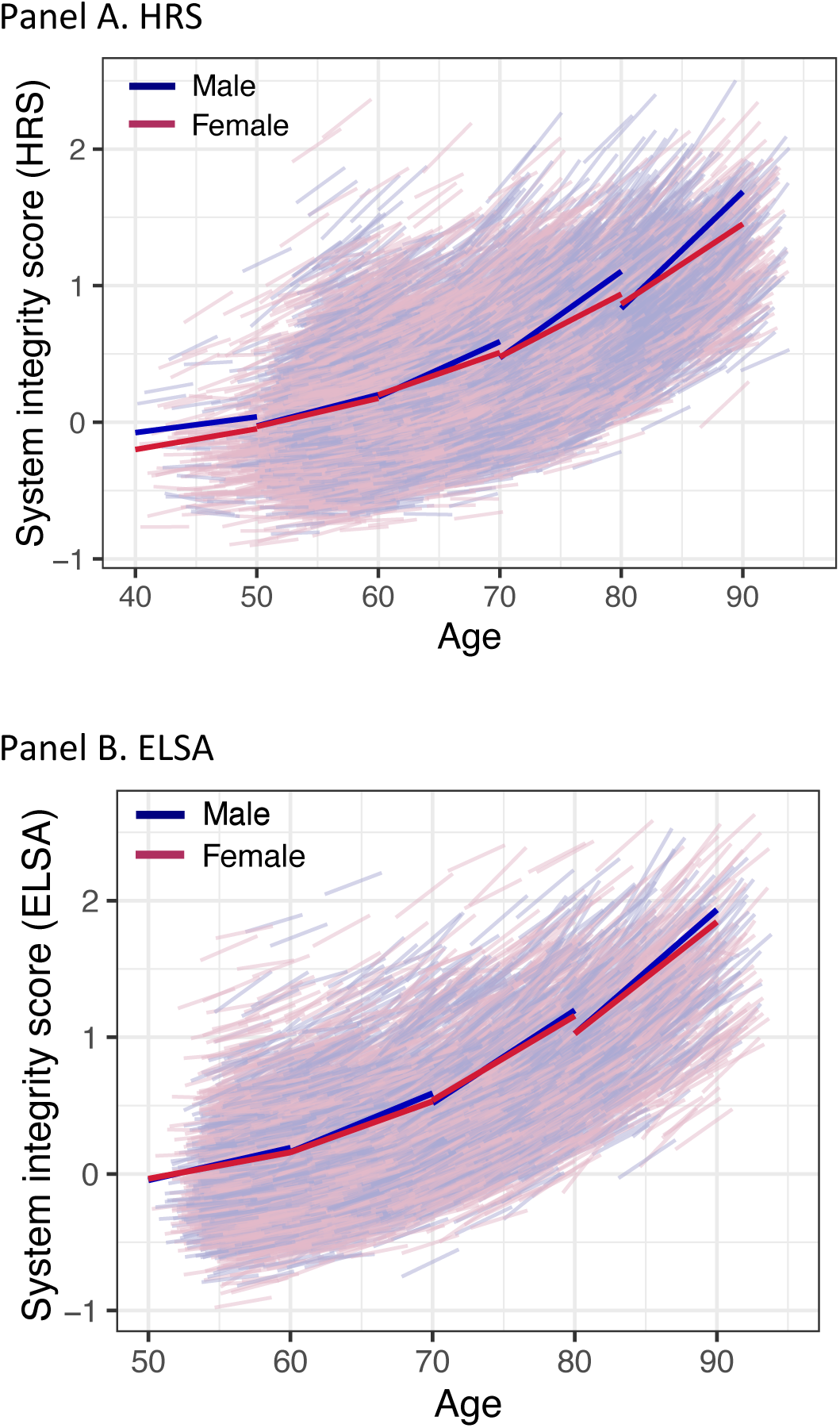
Pace of Aging in the US Health and Retirement Study and the English Longitudinal Study of Aging. Panel A shows data from men and women in the US Health and Retirement Study (HRS, N=13,573). Panel B shows data from men and women in the English Longitudinal Study of Aging (ELSA, N=7,890). The panels show Pace of Aging data for women (pink) and men (blue). Pace of Aging data are graphed as trajectories of system integrity scores. Trajectories were defined from model-predicted intercepts and slopes for biomarkers included in Pace of Aging analysis. Intercepts and slopes were averaged across biomarkers to compute system integrity parameters.

In the United States, in addition to age and sex, healthy aging trajectories differ by race/ethnicity. In HRS, we compared Pace of Aging between participants identifying as White (n=9,301), Black (n=2,269), Hispanic (n=1,674), and Other (n=377) race/ethnicity. Compared to White-identifying participants, Black-and Hispanic identifying participants had faster Pace of Aging (for Black, Cohen’s d=0.18, 95% CI [0.16-0.21]; for Hispanic, Cohen’s d=0.06, 95% CI [0.03-0.10]; **Figure 3; Supplemental Table 6**). Subsequent analyses included participants’ sex, age and, in the HRS, race/ethnicity as covariates.

**Figure 3.**
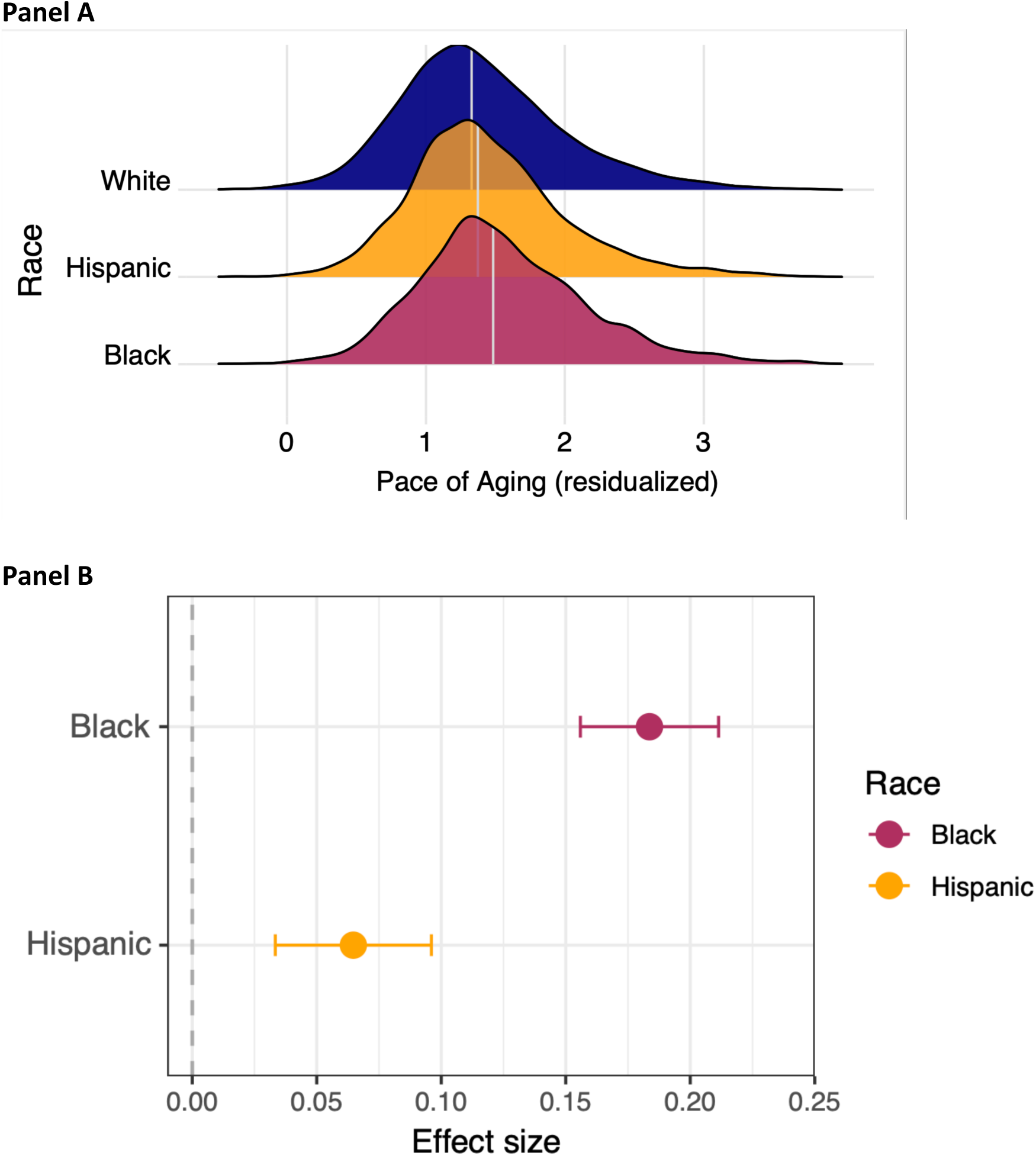
Differences in Pace of Aging among US Older Adults by Race and Ethnicity. Figure shows data from non-Hispanic White (N=9,301), Black (N=2,269), and Hispanic (N=1,674) - identifying older adults in the US Health and Retirement Study. Panel A shows the differences in distribution of Pace of Aging between White, Black, and Hispanic participants. Densities reflect distributions of Pace of Aging after adjustment for chronological age. White lines show group means. Panel B shows effect-size estimates for the differences in Pace of Aging relative to White participants. The figure illustrates overall faster pace of aging in Hispanic- and Black-identifying older adults as compared with White-identifying older adults.

### Participants with faster Pace of Aging were at increased risk of cognitive impairment, incident chronic disease, disability, and mortality

We conducted criterion validity analysis by testing if participants with faster Pace of Aging were more likely to die or more often developed new chronic disease, disability, and cognitive impairment over follow up. We conducted analysis of mortality in HRS using follow-up data accumulated through 2021. We included data on all deaths occurring subsequent to a participants’ second biomarker data collection (i.e. after the minimum follow-up required to compute Pace of Aging). Analysis included N=13,573 participants who contributed mean follow-up time of 10 years (SD=2) over which 3,124 deaths were recorded. Participants with faster Pace of Aging were at increased risk of mortality (HR=1.73 [1.66-1.81], p<0.001; **Figure 4**). We measured chronic disease and disability from participant reports of physician-diagnosed conditions and limitations to activities of daily living (ADLs) and instrumental activities of daily living (IADLs) at baseline and in 2020 (n=8,623). Those with faster Pace of Aging reported more new diagnoses of chronic diseases and more new ADLs and IADLs (chronic diseases IRR=1.08 95% CI [1.06-1.10]; ADLs IRR=1.58 [1.49-1.65]; IADLs 1.59 [1.40-1.66]; all p-values<0.001) and were more likely to develop incident cognitive impairment or dementia (IRR= 1.51 [1.40-1.64]. We conducted parallel analysis of chronic diseases, ADLs and IADLs in ELSA. Although cognitive impairment and dementia classifications were not available in ELSA, we were able to conduct analysis of a cognitive performance score parallel to the one used in HRS. Results were similar. Effect-sizes for all healthspan outcomes are shown in **Figure 5** and **Supplemental Table 7.**

**Figure 4.**
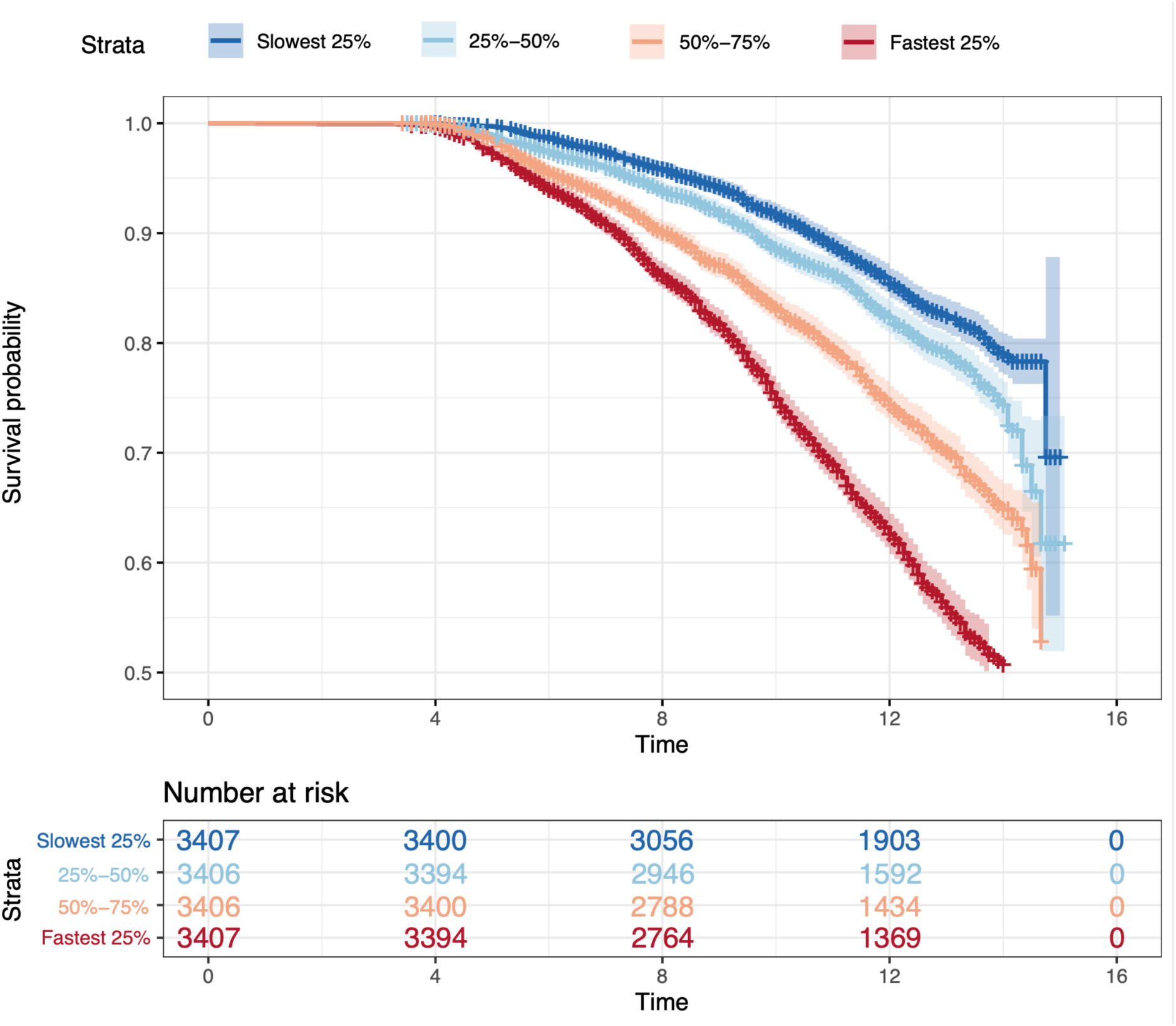
Association of Pace of Aging with mortality. The figure plots Kaplan-Meier curves for four groups of US Health and Retirement Study participants defined by Pace of Aging (slowest quartile graphed in dark blue, fastest quartile graphed in dark red, middle quartiles graphed in lighter shades). Mortality follow-up was conducted from the time of the second biomarker measurement through 2021. Over follow-up there were 1568 deaths in the slowest Pace of Aging quartile, 1990 deaths in the quartile containing the 25^th^-50^th^ percentiles, 1824 deaths in the quartile containing the 50^th^-75^th^ percentiles, and 1946 deaths in the fastest quartile. Numbers at risk at baseline, four, eight, and twelve years of follow-up are shown below the plot.

**Figure 5.**
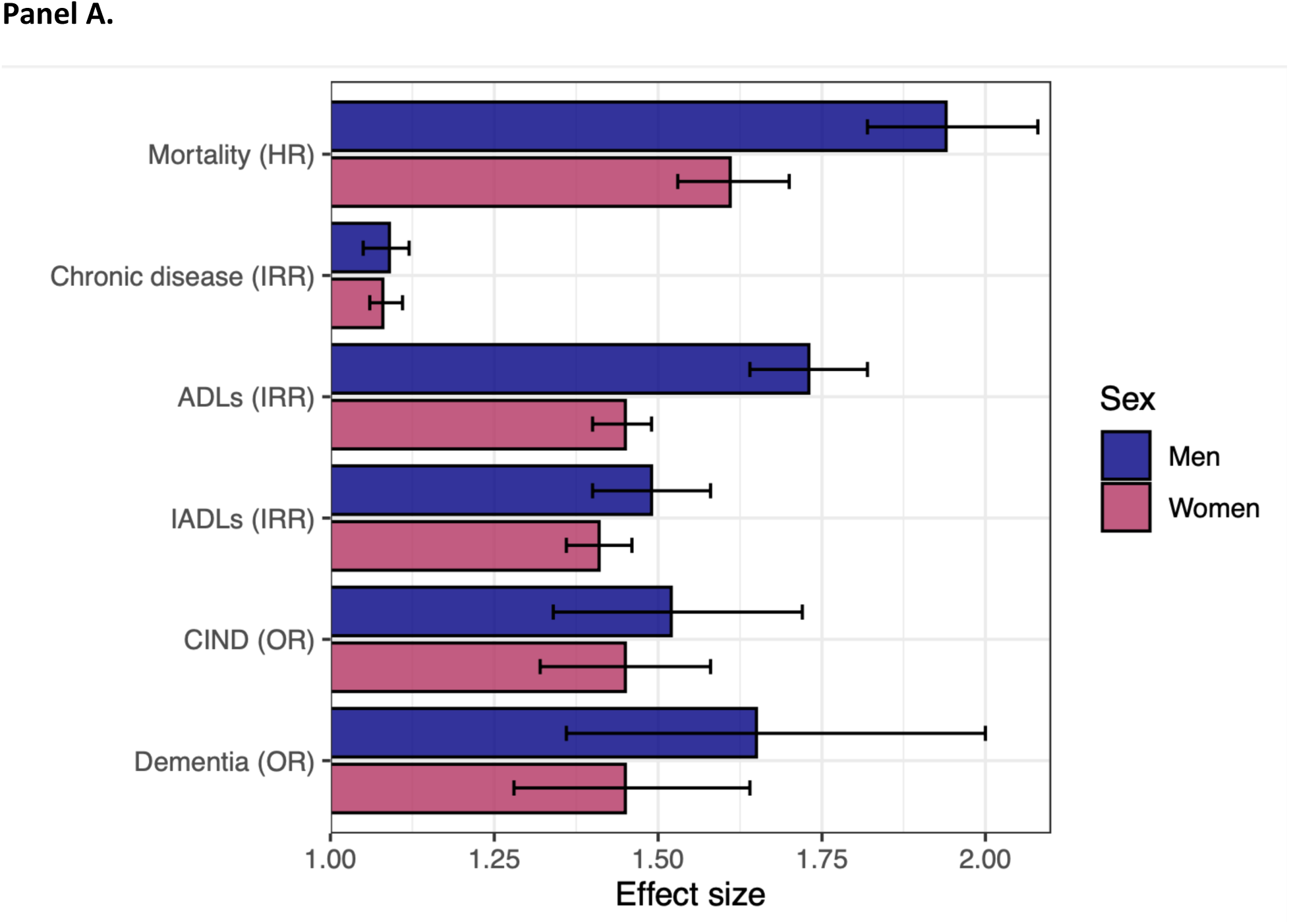
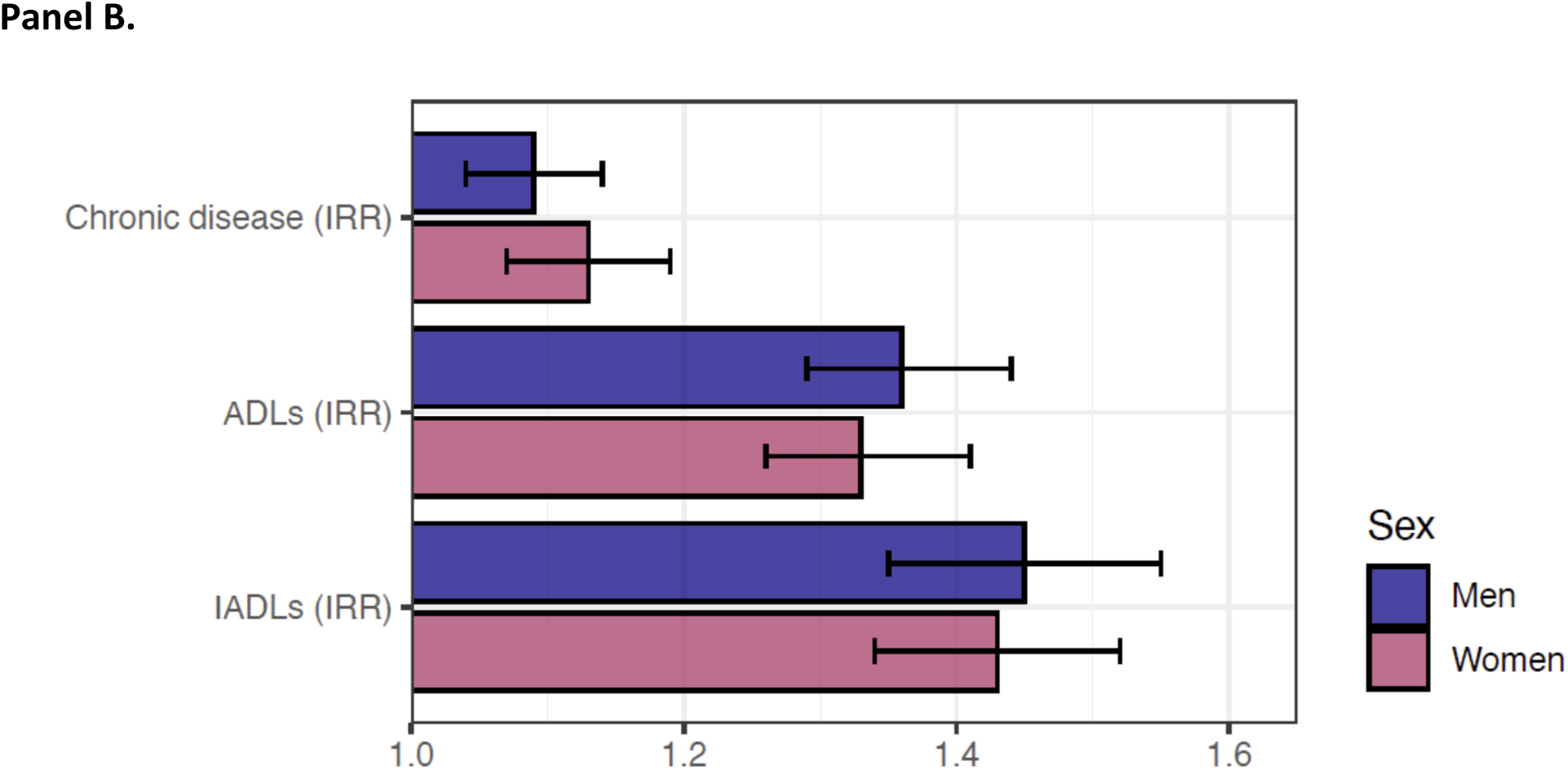
Effect sizes for associations of Pace of Aging with mortality and incident chronic disease, ADLs, IADLs and cognitive impairment. Panel A shows effect-sizes for associations of Pace of Aging with mortality (N=13,573) and with incident chronic disease (n=11,619), ADLs and IADLs (n=11,611), and cognitive impairment (n=10,666) among participants followed-up during 2016-2021 in the US Health and Retirement Study. Panel B shows effect-sizes for associations of Pace of Aging with incident chronic disease, ADLs and IADLs (n=7,872) among participants followed-up during 2014-2018 in the English Longitudinal Study of Aging. Incident chronic disease, ADLs, IADLs, and cognitive impairment were determined by comparing measurements taken at participants’ biomarker baseline wave with measurements taken at the last measurement wave they participated in after Pace of Aging follow-up. Effect-sizes for mortality are reported as hazard ratios (HRs) estimated from Cox regressions. Effect-sizes for incidence of chronic disease, ADLs, and IADLs are reported as incidence rate ratios (IRRs) from Poisson regressions. Effect-sizes for incidence of cognitive impairment are reported as odds ratios (ORs) estimated from multinomial logistic regressions in which outcomes were normal (reference), cognitively impaired but not demented (CIND), and demented. Effect-sizes are reported for a one-standard-deviation difference in Pace of Aging. All regression models included covariate adjustment for sex, race, age, age-squared, a set of terms encoding baseline measurement year for Pace of Aging, follow-up time to outcome assessment, and whether the participant was aged 65 or older at baseline.

To evaluate Pace of Aging in the context of existing approaches to quantification of biological aging, we conducted analysis comparing pace of Aging with three published metrics of biological age in HRS. We analyzed versions of the Homeostatic Disregulation (HD)^22^, Klemera-Doubal method Biological Age (KDM) ^23^, and PhenoAge ^24^ algorithms adapted for the HRS in previous work ^25,26^. In contrast to Pace of Aging, these three metrics are implemented in a single cross-section of data and attempt to represent the progress of aging rather than its current rate. We conducted three analyses. First, we calculated correlations among the measures. After residualization for chronological age, the three biological age metrics correlated with Pace of Aging at r=0.3-0.4. Second, we compared effect sizes with healthspan outcomes. Across healthspan outcomes, effect sizes for Pace of Aging were larger than effect sizes for the other metrics, with the exception of chronic disease incidence, for which Pace of Aging effect sizes and PhenoAge effect sizes were similar. Third, we tested independence of Pace of Aging associations with healthspan outcomes from the biological age metrics. Across healthspan outcomes, Pace of Aging associations were only modestly attenuated after covariate adjustment for biological age metrics and remained statistically different from zero at the p<0.05 level. The largest attenuation was for analysis of mortality. After adjustment for

PhenoAge, the effect size for Pace of Aging was reduced to HR=1.43 [1.31-1.56]. Results from comparative analysis are shown in **Supplemental Figure 8.**

To evaluate the sensitivity of Pace of Aging associations with healthspan outcomes to the specific biomarker composition of the measure, we performed leave-one-out analysis. In this analysis, we composed Pace of Aging from subsets of eight of the total nine biomarkers and re-estimated associations with healthspan outcomes. Results were consistent across leave-one-out specifications, with the exception that, in ELSA, associations with cognitive decline were reduced to near zero when balance-test data were removed (**Supplemental Figure 7**).

We observed some differences in effect-sizes for associations of Pace of Aging with healthspan outcomes between demographic groups of participants. Effect-sizes were generally somewhat larger for white participants, men, and those under age 65 as compared with non-white participants, women, and older participants. Results are reported in **Supplemental Tables 7** and **8** and **Supplemental Figures 6** and **7**.

### Pace of Aging associations with cognitive impairment, morbidity, disability, and mortality were independent of smoking, obesity, and education

We investigated whether Pace of Aging associations with cognitive impairment, morbidity, disability, and mortality were accounted for by baseline socio-economic and health behavior risk factors. HRS participants with lower levels of education, who were overweight or obese, and who were current or former smokers tended to have a faster Pace of Aging (Cohen’s d range 0.08 – 0.38). We therefore repeated analyses of cognitive function and impairment, morbidity, disability, and mortality including these risk factors as covariates. Adjusted results were similar to unadjusted results. Results were similar in ELSA, with the exception that covariate adjustment for BMI attenuated association with cognitive function below the level of statistical significance. Full results are reported in **Supplemental Tables 7** and **8**.

## DISCUSSION

The Pace of Aging method remains an understudied approach to quantification of biological aging. The method was first proposed in the Dunedin Longitudinal Study^16^, which has followed a single-year birth cohort over five decades^27^. The Dunedin analysis included individuals all born in the same year and followed-up at the same ages from young adulthood to midlife. Here, we show that Pace of Aging can be modeled in the very different setting of a national-population-representative cohort of older adults followed-up at ages ranging from the sixth through ninth decades of life. This contribution advances translation of the geroscience hypothesis into the domain of population science. Omics-based measurements are not yet available in most national cohorts or, as in the case of the HRS, are available for only a select subsample of participants. Existing methods for quantification of biological aging from data routinely collected in national studies measure the progress of aging, not its current rate. The adapted Pace of Aging method introduced here allows for measurement of the aging rate in the context of national cohorts, providing planners and policy makers with a new tool to understand population aging and promote healthy longevity. In addition, we report three key findings that advance knowledge of Pace of Aging.

First, Pace of Aging accelerates at more advanced ages. HRS participants who were older at their baseline biomarker assessment showed more rapid change across subsequent follow-ups as compared to those who were younger. This observation is consistent with biodemographic data showing that mortality risk accelerates at older ages^28^, with biomarker analysis in the Baltimore Longitudinal Study on Aging^29^ and the Cardiovascular Health Study^30^, and with analysis of the Pace of Aging epigenetic clock, DunedinPACE^16,31^, and advances the hypothesis that the pace of aging accelerates later in life.

Second, Pace of Aging is faster in sociodemographic groups characterized by shorter lifespan. Men tended to experience faster Pace of Aging as compared with women. Those with less education tended to experience faster Pace of Aging as compared to those with more education, consistent with observations of a socioeconomic gradient in the pace of aging from the Dunedin Cohort and a Swiss cohort^32,33^. In addition, we observed a faster Pace of Aging in Black- and Hispanic-identifying as compared to White-identifying Americans. While race/ethnic disparities in healthy aging outcomes are well established^34^, our observation of faster Pace of Aging in Black as compared to White Americans indicates healthy-aging disparities continue to accumulate into later life. This finding suggests that intervention/prevention in this older-aged group has the potential to reduce aging-related health disparities.

Third, midlife and older adults with faster Pace of Aging were at increased risk of incident chronic disease, disability, and mortality. In the Dunedin Study, where we first introduced Pace of Aging, participants are still middle aged; it is not yet possible to test associations with aging-related health problems and mortality. In HRS, older adults with faster Pace of Aging more often developed new chronic diseases and disabilities and were at increased risk of death.

Moreover, these associations were independent of smoking, obesity, and educational attainment. These findings contribute evidence that the adapted Pace of Aging method introduced here captures differences in aging processes that are important to healthspan and lifespan. The variation observed across social and demographic groups suggests potential to modify pace of aging through changes in the organization of aging societies and in the environment and behavior of older adults.

These findings have implications for future research. Pace of Aging summarizes changes occurring across multiple systems in the body to provide a dynamic measure of a person’s healthy aging trajectory. The availability of a Pace of Aging measure within the HRS can inform research in the fields of Sociology and Economics to understand how social transitions such as retirement, spousal loss, and caregiving responsibilities affect trajectories of healthy aging. In medicine and gerontology, Pace of Aging measures can help reveal how health shocks such as new diagnoses, accidents, or elective surgeries modify aging trajectories. In life-course epidemiology, Pace of Aging measures can help differentiate effects of early-life exposures on aging-related health decline from health deficits established earlier in life. In health equity research, Pace of Aging measures can be used to quantify disparities across population subgroups in health aging trajectories and illuminate how economic and health shocks contribute to inequalities^35^. These lines of research, in turn, can deliver new knowledge critical to the development of policies and programs to promote healthy longevity, consistent with the priorities of World Health Organization^14^ and other public health actors^36–38^.

We acknowledge limitations. The HRS measurement battery available to measure Pace of Aging is more limited as compared with the Dunedin Study. Some parameters are measured with lower precision instruments (e.g. peak flow meters as compared to spirometry for assessment of lung function). In other cases, measurements available the in Dunedin Study were not collected by HRS (e.g. periodontal health, cardiorespiratory fitness). In addition, the older ages of HRS participants limits utility of some biomarkers that are available. For example, medical management of systolic blood pressure and cholesterol levels in older adults along with non-linearities in the patterning of these markers with aging in late life limit their utility to Pace of Aging analysis. Nevertheless, HRS data do provide for measurement of changes in pulmonary, vascular, immune, renal, endocrine, metabolic, and musculoskeletal systems and therefore make possible Pace of Aging analysis in a large, national sample of older adults with significant race/ethnic diversity.

HRS biomarker data have been collected at up to only three timepoints for each participant. As a result, we were unable to model non-linear changes in biomarkers. Consequently, we must infer the acceleration in Pace of Aging at older chronological ages from comparisons of younger and older participants. As additional waves of biomarker data are collected, it will be possible to model non-linear slopes of change and more rigorously test the hypothesis that aging accelerates towards the end of life.

Because HRS enrolls some participants in their 70s and 80s, the sample we analyzed may over-represent those with slower Pace of Aging. If, as data reported here suggest, faster Pace of Aging increases risk of death, the oldest segments of the sample will tend to include those with slower Pace of Aging. Importantly, despite this limitation, we still observe evidence of faster Pace of Aging in older as compared to younger participants. Moreover, faster Pace of Aging was associated with incident morbidity, disability, cognitive impairment, and mortality, in both older and younger segments of the sample. Further analysis in samples with large numbers of adults aged 80-plus followed over multiple time points is needed to establish utility of the Pace of Aging method in this population.

Beyond the potential bias toward slower agers, our analysis sample was limited to participants contributing at least two repeated observations of biomarker data, requiring follow-up over at least four years. The subset of HRS participants meeting this requirement tended to be better educated, less diverse, and younger than the overall sample. This may also contribute to the observed distribution of the Pace of Aging being somewhat slower than in the general population HRS is designed to represent.

Consistent with observations in the original Pace of Aging analyses^15,39^, we found that patterns of decline were correlated across biological systems although, as in the original analyses, these correlations were modest. Other investigators have interpreted similarly modest cross-system correlations as evidence of stochastic variation in aging^40,41^. Whether Pace of Aging reflects an underlying set of biological processes causing correlated change across systems or instead summarizes changes across systems experiencing stochastic aging trajectories is an important question for further research. Critically, either interpretation supports application of Pace of Aging within population health science to quantify trajectories of healthy aging.

To summarize, we developed a measure of Pace of Aging in HRS. It is predictive of morbidity, disability, cognitive impairment, and mortality and reflects known social gradients in healthy aging. It provides a new tool for researchers seeking to understand how features of societal organization, built and social environments, and individual behavior contribute to healthy aging trajectories in populations around the world.

## Data Availability

All data produced are available online at https://hrsdata.isr.umich.edu/data-products

